# Genomic Surveillance and Evolutionary Dynamics of Influenza A Virus in Sri Lanka

**DOI:** 10.1101/2024.08.23.24312476

**Authors:** Tibutius Thanesh Pramanayagam Jayadas, Chandima Jeewandara, Bhagya Senadheera, Heshan Kuruppu, Rivindu Wickramanayake, Farha Bary, Ananda Wijewickrama, Suranga Manilgama, Manouri Gamage, Nilanka Perera, Graham S. Ogg, Gathsaurie Neelika Malavige

## Abstract

**Background:** Influenza A has been named as a priority pathogen by the WHO due to the potential to cause pandemics. Genomic sequencing of influenza strains is important to understand the evolution of the influenza strains and also to select the appropriate influenza vaccines to be used in the different influenza seasons in Sri Lanka. Therefore, we sought to understand the molecular epidemiology of the influenza viruses in the Western Province of Sri Lanka, including mutational analysis to investigate the evolutionary dynamics.

**Methodology:** A total of 349 individuals presenting with fever and respiratory symptoms were enrolled in this study from November 2022 to May 2024. Nasopharyngeal and oropharyngeal specimens were collected and screened using quantitative PCR to detect Influenza A, Influenza B, and SARS-CoV-2. Subtyping and genomic sequencing was carried out on influenza A strains using Oxford Nanopore Technology.

**Results:** Influenza A was detected in 49 (14 %) patients, influenza B in 20 (5.7%) and SARS-CoV-2 in 41 (11.7%). Co-infections were observed in five participants. The phylogenetic analysis assigned the H1N1 HA gene sequences within the 6B.1A.5a.2a clade. The HA gene of the H1N1 sequences in 2023 were assigned as belonging to the subclades C.1, C.1.2, and C.1.8, while the 2024 sequences were assigned to subclades C.1.8 and C.1.9. The H3N2 sequences from 2023 were assigned to the 3C.2a1b.2a.2a.1b clade and subclade G.1.1.2, while the 2024 sequences were assigned to the 3C.2a1b.2a.2a.3a.1 clade and subclade J.2. The K54Q, A186T, Q189E, E224A, R259K, K308R, I418V, and X215A amino acid substitutions were seen in the H1N1 in the 2023 and 2024 sequences. The 2024 H1N1 sequences additionally exhibited further substitutions, such as V47I, I96T, T120A, A139D, G339X, K156X, and T278S.

**Conclusion:** In this first study using genomic sequencing to characterize the influenza A strains in Sri Lanka, which showed different influenza A viruses circulating in an 18-month period. As the Sri Lankan strains also had certain mutations of unknown significance, it would be important to continue detailed surveillance of the influenza strains in Sri Lanka to choose the most suitable vaccines for the population and the timing of vaccine administration.

## Introduction

Seasonal influenza outbreaks are associated with significant morbidity and mortality, with estimated cases 3.2 million cases of severe disease each year, globally [1]. Due to the potential of influenza A strains causing pandemics, it has been included in the WHO pathogen prioritization list published in 2024 [2]. Despite the availability of effective vaccines and antivirals, the WHO estimates that 290,000 to 650,000 deaths occur annually due to this virus [3]. Those at extremes of age, pregnant women, individuals with comorbidities and immunocompromised individuals are at risk of developing severe disease and death [1]. Due to the rapid evolution of the virus and emergence of avian influenza in certain regions in the world, genomic surveillance of influenza strains in crucial to monitor the influenza strains that cause outbreaks in different countries.

Influenza viruses belong to the Orthomyxoviridae family and are classified into four types: A, B, C, and D. Among these types, the influenza A virus has been responsible for several major pandemics in the past century, including the 1918 Spanish flu (H1N1), the 1957 Asian flu (H2N2), and the 1968 Hong Kong flu (H3N2), all of which caused a significant global health burden [4]. Influenza A viruses are categorized based on the properties of their surface glycoproteins, hemagglutinin (HA) and neuraminidase (NA) [5]. The influenza A virus is classified into different subtypes based on the HA and NA glycoproteins. There are 18 known HA subtypes (H1 to H18) and 11 known NA subtypes (N1 to N11) [5]. The interplay of antigenic shift and drift among these subtypes results in generation of multiple strains due to varied combination of HA and NA subtypes [6]. Reassortment events, facilitated by natural reservoirs such as swine, birds, and horses, contribute to the emergence of novel strains with pandemic potential [6].

Globally, influenza A continues to exhibit seasonal patterns, with peaks typically occurring during the winter months in temperate regions and outbreaks often coinciding with the monsoon season in tropical and subtropical regions [7]. Although vaccination has been proven to be effective in providing some protection against influenza, they need to be given annually due to the changes in the circulating strains of the virus [8]. Therefore, the WHO Global Influenza Program recommends an evidence-based approach by grouping countries with similar seasonality patterns and virus antigenic characteristics into Influenza Vaccination Zones to address specific country needs [9]. The WHO encourages countries to conduct local surveillance to assess their seasonality patterns and circulating strains to facilitate the decision on selecting Northern Hemisphere (NH) and Southern Hemisphere (SH) vaccines and to determine the timing of vaccination campaigns [9].

Sri Lanka is a tropical country and influenza viruses circulate throughout the year, with two peaks typically occurring during the rainy seasons which are from May to July and from November to January[10]. As a part of the integrated SARS-CoV-2 and influenza surveillance platform, limited number of samples are subjected to testing for the presence of influenza A and SARS-CoV-2, which are then subjected to subtyping if influenza A is identified. However, genomic sequencing is not carried out, which is important to identify the origin and evolution of the influenza strains and also to select the appropriate influenza vaccines to be used in the different influenza seasons in Sri Lanka. In this study, we carried out proceed to understand the molecular epidemiology of the influenza viruses in the Western Province of Sri Lanka, including mutational analysis to investigate the evolutionary dynamics.

## Methodology

### Recruitment of patients and collection of samples

Nasopharyngeal and oropharyngeal specimens were collected from both 349 adult and paediatric patients presenting with an acute febrile illness with respiratory symptoms such as cough, sore throat, and rhinorrhea. Patients were recruited from two tertiary care hospitals, which were the National Institute of Infectious Disease and Colombo South Teaching Hospital, situated in the Western Province of Sri Lanka, between November 2022 to May 2024. Patients with a duration of illness of 7days were included in the study.

### Ethics approval

Informed written consent was taken from all adult patients and in the case of paediatric patients, informed written consent was obtained from their parents/guardian. Ethics approval was obtained from the Ethics Review Committee, University of Sri Jayewardenepura.

### Screening for Respiratory Viruses Using Quantitative Polymerase Chain Reaction (qPCR)

Viral RNA was extracted using Applied Biosystems™ MagMAX™ Viral/Pathogen Nucleic Acid Isolation Kit. All the collected samples were screened for Influenza A, Influenza B, using Respiratory Panel 1 qPCR Kit and when influenza A virus was detected it was also subtyped using the Viasure, Spain (VS-RPA112L v.03). Concurrently, each sample was tested for the presence of SARS-CoV-2 using TaqPath™ COVID-19 CE-IVD RT-PCR Kit (Thermo Fisher Scientific, USA).

### Library preparation and sequencing of the influenza A virus

All 49 samples that were identified as being infected with influenza A by qPCR were chosen for ONT sequencing. The reaction mixture was prepared using 12.5 µL of Superscript III One-Step PCR reaction buffer, 0.5 µL of SuperScript III RT/Platinum Taq Mix (Thermo Fisher Scientific, USA), and primers (MBTuni-12 at 0.1 µM, MBTuni-12.4 at 0.1 µM, and MBTuni-13 at 0.2 µM). Additionally, 2.5 µL of RNA template was added, and PCR grade water was used to attain a final volume of 25 µL [11]. The PCR reactions were carried out with an initial incubation at 42 °C for 60 minutes, followed by denaturation at 94 °C for 2 minutes. This was succeeded by 5 cycles of denaturation at 94 °C for 30 seconds, annealing at 45 °C for 30 seconds, and extension at 68 °C for 3 minutes. Subsequently, 20 cycles were performed with denaturation at 94 °C for 30 seconds, annealing at 58 °C for 30 seconds, and extension at 68 °C for 3 minutes, concluding with a final extension at 68 °C for 10 minutes.

Libraries for sequencing were generated from the amplified samples using the ONT Rapid Barcoding Kit (SQK-RBK110.96), following the protocol version RBK_9126_v110_revO_24Mar2021. The pooled barcoded MinION library was subsequently loaded onto the MinION Mk1b sequencer from Oxford Nanopore Technologies, Oxford, United

Kingdom, equipped with an R9.4 flow cell. Real-time base calling was performed using MinKNOW version 3.0.4 with the Guppy base calling software version 3.2.10.

### Generation of Consensus sequences (EPI2ME)

Base calling was performed using the Guppy (version 6.5.7) with Fast model, 450 bps base calling model. The resulting reads were analyzed using the wf-flu workflow. Samples that were unclassified were excluded from further analysis. All samples that were successfully classified as Archetypes were subsequently submitted to the GISAID database.

### Construction of phylogenetic trees

We used HA and NA genes sequences with >90% coverage to create the phylogenetic trees. Accession numbers included in the analysis are included in Supplementary Table.1. From 2021 to 2024 sequences from WHO South-East Asian region (100 simple random samples), WHO global (100 random samples) and vaccine reference sequences in GISAID database was used to construct the phylogenetics trees for HA gene and NA gene. Phylogenetic analyzes for all IAV segments were performed. Sequence alignments were separately constructed for HA (H1 and H3 subtypes), NA (N1 and N2 subtypes). 222 sequences were included for analysis of H1, 209 for H3, 224 for N1 and 210 sequences for N2.

Multiple sequence alignment was generated using MAFFT v.7.508 employing the FFT-NS-i algorithm. Subsequently, this multiple sequence alignment was used to infer a Randomized Axelerated Maximum Likelihood (RAxML) phylogenetic tree using RAxML (v.8.2.12) with GTRGAMMA substitution model and bootstrap of 1000 replicates. The best-fit model GTR+F+R5 was chosen using ModelFinder. Final visualizations of the phylogenetic tree were done using R\ggtree, R\ape and R\ggstar packages (R version 4.1.2).

### Mutational analysis

Mutation analysis was carried out for the sequenced samples, prior to the variant calling, by removing the signal peptides in the H1 and H3 genes. To identify mutations in the H1N1 sequences, they were compared with the A/Wisconsin/588/2019 strain (EPI_ISL_19085699), which is the 2021-2022 Northern Hemisphere vaccine strain for H1NI [12]. To identify mutations in the Sri Lankan H3N2 sequences, they were compared with the A/Darwin/6/2021 (EPI_ISL_1563628) which was the 2022 Southern Hemisphere vaccine strain for H3N2[13]. The predicted position of the signal peptide in the sequences were identified with SignalP-5.0 tool. Based on the analysis using this predictive model, we identified that the predicted position of the signal peptide in the A/Wisconsin/588/2019 strain was in the positions in the amino acid positions, 1 to 17 (likelihood ratio 0.797) and for the A/Darwin/6/2021, amino acid positions 1 to 16 (likelihood ratio, 0.6971). Mutations were analyzed and visualized with R packages (R version 4.1.2) after removing the signal peptide region from the sequence of the protein.

## Results

Of the 349 patients recruited in the study, 173 (49.5%) were males and 176 (50.4%) were females and 205 (58.7%) were adults. Influenza A was detected in 49 (14 %) patients, influenza B in 20 (5.7%) and SARS-CoV-2 in 41 (11.7%). Co-infections were observed in five participants: four were co-infected with both Influenza A and B, and one individual was co-infected with Influenza A and SARS-CoV-2. The age distribution of these infections in different age groups is shown in figure 1. Notably, the highest incidence of influenza A (42.8%) and influenza B (5.7%), was detected in children <10 years of age. In contrast, the highest incidence of SARS-CoV-2 infection was seen in individuals > 60 years old, with 22.7% of the infections been detected in this age group, while 6/66 (9.1%) were infected with influenza A.

**Figure. 1:**
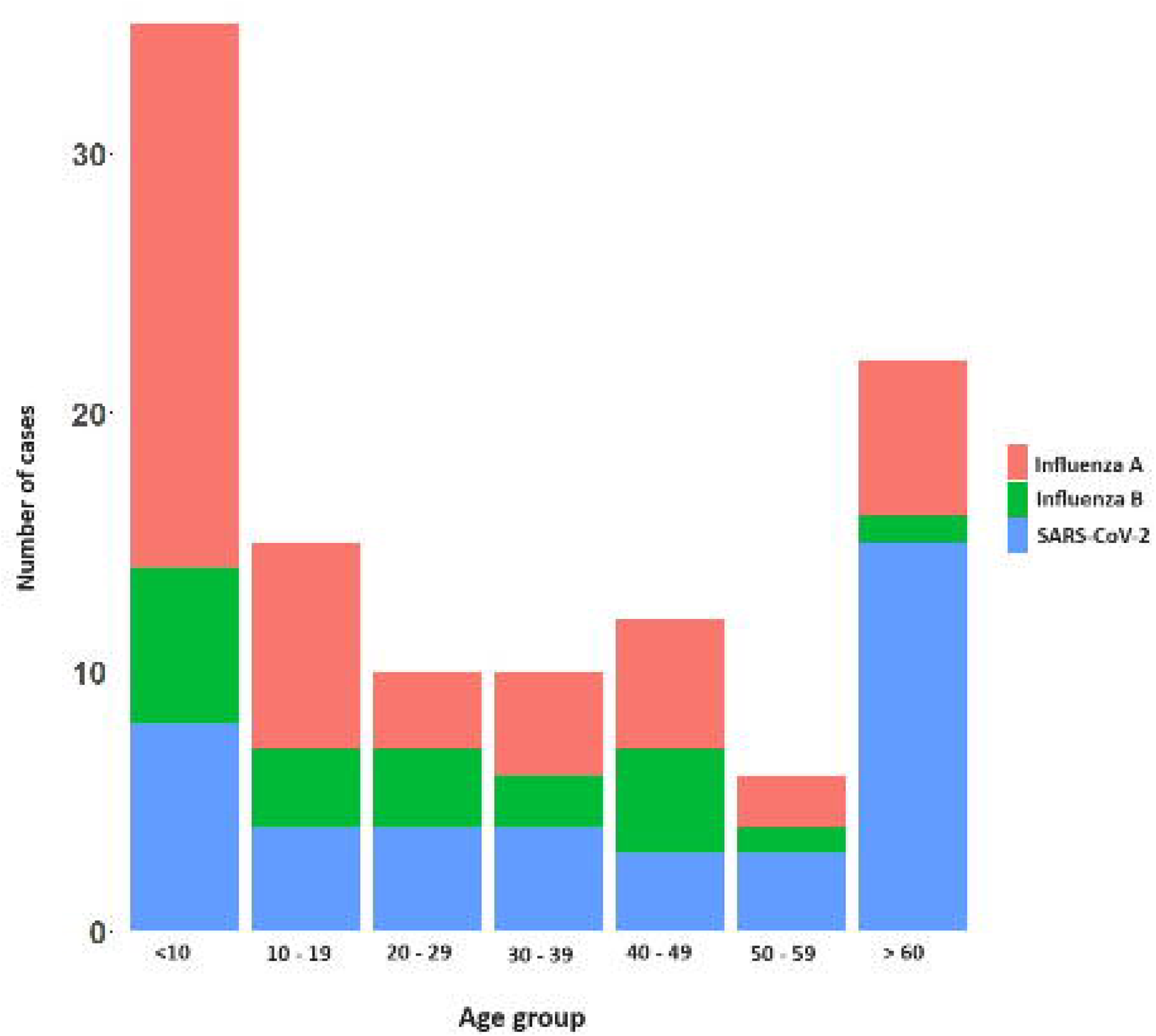
Distribution of influenza A, influenza B, and SARS-CoV-2 infections across different age groups among recruited participants

### Influenza A virus subtyping, and seasonal patterns of infection

Infection due to influenza A, influenza B and SARS-CoV-2 was detected during the study period of November 2022 to May 2024. We paused the study during the months of July to October 2023, where very limited cases of respiratory infections were reported in both tertiary care hospitals. Of the individuals who tested positive for Influenza A, 23 identified as H1N1, 18 as H3N2, while 8 infections could not be classified. From December 2022 to February 2023, H1N1 was the predominant subtype of Influenza A (Supplementary Figure 1). However, a significant shift occurred from early March 2023 to July 2023, with H3N2 becoming the dominant strain. By December 2023, a resurgence of the H1N1 subtype was observed.

### Phylogenetic Analysis of H1N1 viruses

Out of the 49 influenza A samples, 21 were successfully sequenced, comprising 17 H1N1 samples and 4 H3N2 samples. Based on sequence quality, 14 HA genes and all 17 NA genes from the H1N1 viruses were included in the phylogenetic analysis, while 3 HA genes and 4 NA genes from the H3N2 viruses were analyzed. The phylogenetic analysis assigned the H1N1 HA gene sequences within the 6B.1A.5a.2a clade. The HA gene of the H1N1 sequences in 2023 were assigned as belonging to the subclades C.1, C.1.2, and C.1.8, while the 2024 sequences were assigned to subclades C.1.8 and C.1.9. Phylogenetic analysis of the H1N1 HA gene revealed that the 2023 sequences were most closely related to strains from Bangladesh and Bangkok, whereas the 2024 sequences were most similar to those from the Maldives (Figure 2).

**Figure 2:**
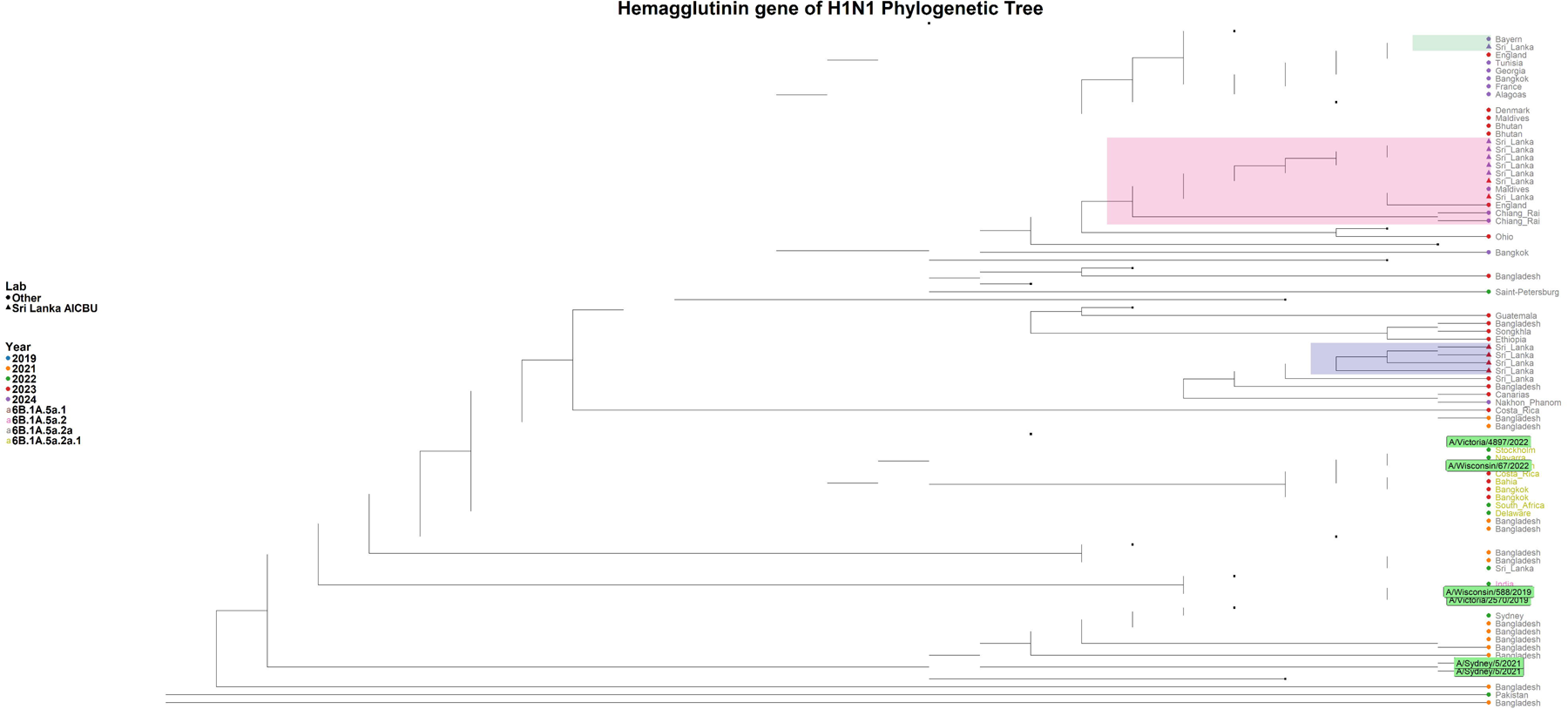
Phylogenetic tree of the H1N1 HA gene. The phylogenetic tree was generated with the Sri Lankan H1N1 sequences (n=14) in comparison to the global H1N1 strains. All the Sri Lankan were assigned to clade 6B.1A.5a.2a. The H1N1 Sri Lankan sequence clusters are shaded in green, orange and grey shades, while the reference sequences are highlighted in green.

The Sri Lankan H1N1 HA gene sequences and the A/Sydney/5/2021 Southern Hemisphere vaccine strain (used in the 2023 Southern influenza vaccine) belong to clade 6B.1A.5a.2a. Although A/Wisconsin/67/2022 and A/Victoria/4897/2022 from the Northern Hemisphere vaccine reference are in the 6B.1A.5a.2a.1 clade, the Sri Lankan HA gene sequences from 2023 and 2024 were more closely related to these Northern Hemisphere H1N1 strains than to the A/Sydney/5/2021 strain. The NA gene analysis showed that 2023 sequence showed close resemblance to the sequences from England and 2024 NA gene sequences closely resembled with sequences from Belgium and Bangladesh (Supplementary Figure 2).

### Phylogenetic Analysis of H3N2 viruses

The H3N2 sequences from 2023 were assigned to the 3C.2a1b.2a.2a.1b clade and subclade G.1.1.2, while the 2024 sequences were assigned to the 3C.2a1b.2a.2a.3a.1 clade and subclade J.2. HA gene analysis revealed that the 2023 sequences were closely related to those from Bangkok (Thailand), Cantabria (Spain), and England, whereas the 2024 sequences show similarity to those from Belgium and Nakhon Pathom (Thailand) (Figure 3). The 2023 HA gene sequence is more closely related to the A/Darwin/6/2021 vaccine strain, while the 2024 HA gene sequence is more similar to A/Massachusetts/18/2022, both of which fall within the 3C.2a1b.2a.2a.3a.1 clade. NA gene analysis of the 2023 H3N2 samples indicated a close relationship with sequences from Catalonia (Spain), Rhode Island (USA) and England, while the 2024 samples showed the similarity to sequences from France (Supplementary Figure 3).

**Figure 3:**
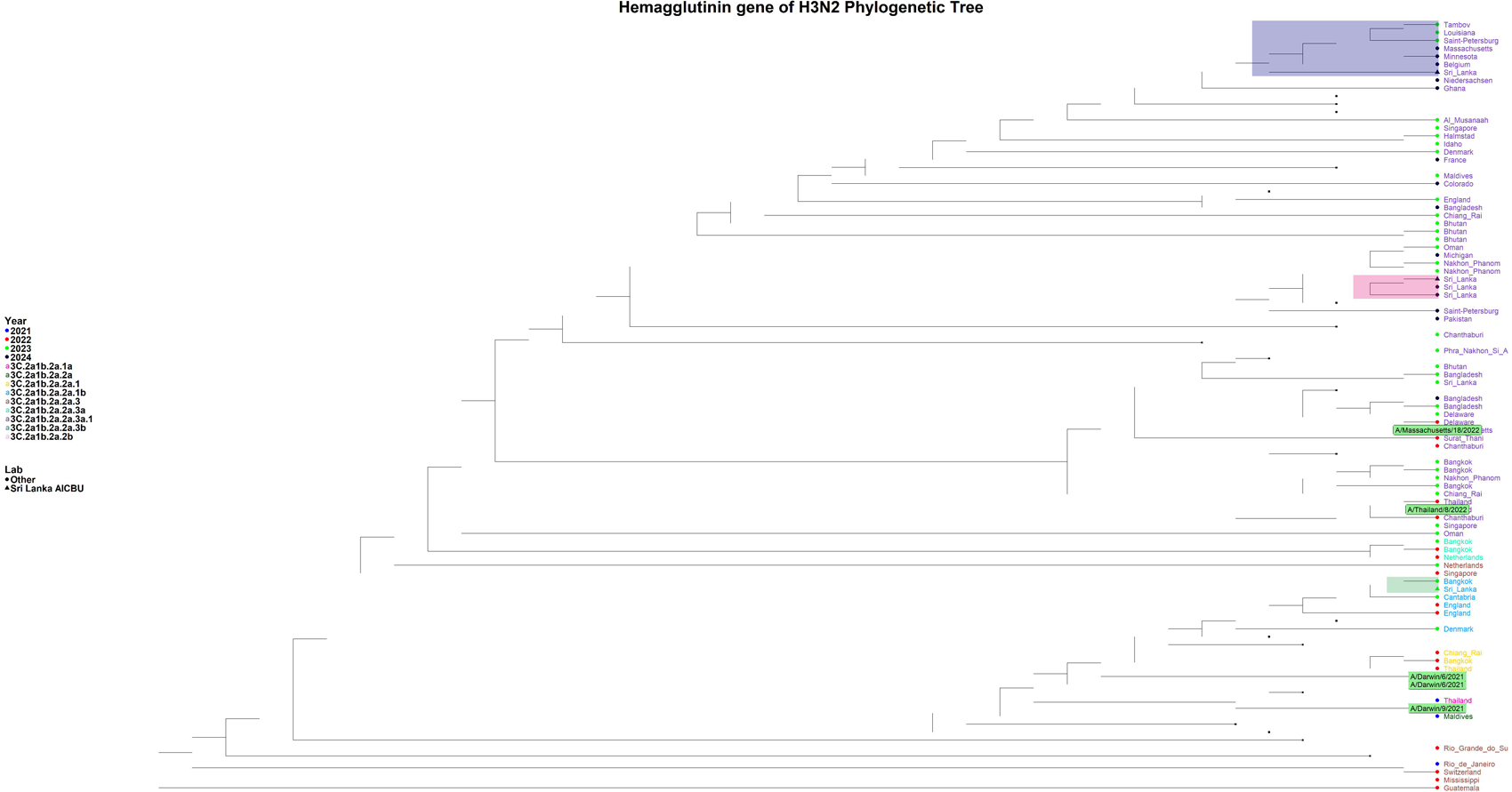
Phylogenetic tree of the H3N2 HA gene. The phylogenetic tree was generated with the Sri Lankan H3N2 sequences (n=3) in comparison to the global H1N1 strains. The 2023 sequence was assigned to the 3C.2a1b.2a.2a.1b clade, while the 2024 sequences were assigned to the 3C.2a1b.2a.2a.3a.1 clade. The H1N1 Sri Lankan sequence clusters are shaded in green, orange and grey shades, while the reference sequences are highlighted in green.

### Mutation analysis

The mutational analysis of the H1N1 hemagglutinin (HA) gene was carried out in reference to the A/Wisconsin/588/2019 (H1N1) strain. Accordingly, we identified amino acid substitutions, including K54Q, A186T, Q189E, E224A, R259K, K308R, I418V, and X215A across both the 2023 and 2024 sequences (Figure 4A). The 2024 H1N1 sequences additionally exhibited further substitutions, such as V47I, I96T, T120A, A139D, G339X, K156X, and T278S. The positions of these mutations and the function of these genes are shown in supplementary table 2. In the neuraminidase (NA) gene, H1N1 sequences identified in 2023 and 2024 shared the X136Q and V453M/V453T substitutions, with the 2024 sequences uniquely showing mutations at I264T, E433X, and E433K (Figure 4B). In comparison to the A/Wisconsin/67/2022 vaccine reference sequence, the HA gene of Sri Lankan H1N1 strains in 2023 and 2024, demonstrated substitutions including S137P, R142K, E260D, A277T, and D356T. The 2024 sequences also presented additional mutations, namely V47I, I96T, and T120A (Supplementary Figure 4A). In the NA gene, the 2024 sequences revealed further substitutions at V13I, S200N, and L339S (Supplementary Figure 4B).

**Figure 4:**
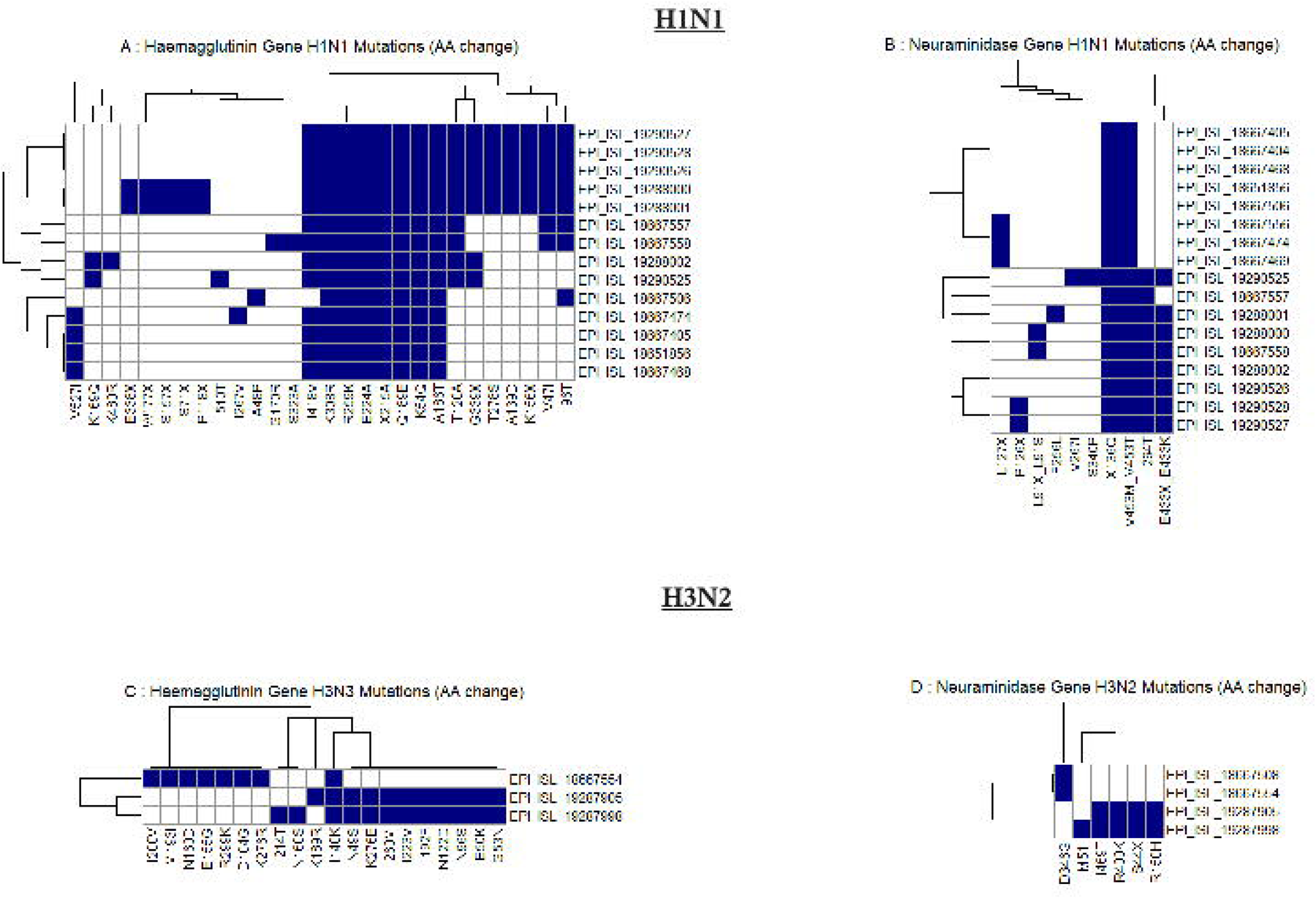
Heatmap of amino acid substitutions in hemagglutinin (HA) and neuraminidase (NA) genes of influenza A H1N1 and H3N2 viruses. (Panels A and B show mutations in the HA and NA genes of H1N1, respectively, while panels C and D display mutations in the HA and NA genes of H3N2. Each row represents an individual virus sequence, identified by its GISAID EPI_ISL accession number, and each column represents a specific amino acid position where mutations have occurred. The presence of a mutation is indicated by a blue square, and the absence by a white square).

In the hemagglutinin (HA) gene of H3N2, using A/Darwin/6/2021 (H3N2) as the reference, the 2023 sequence revealed several amino acid substitutions, including I200V, M193I, N160D, E155G, R299K, D104G, and K276R. The I140K substitution was consistently observed across all analyzed sequences. In contrast, the 2024 sequences exhibited additional substitutions such as K189R, N49S, K276E, I260M, I223V, I192F, N122D, N96S, E50K, and G53N (Figure 4C). Due to the E50K and I223V substitutions, our H3N2 strains in 2024, are most similar to the A/Thailand/8/2022, subclade J). In comparison to the A/Massachusetts/18/2022 vaccine strain, both the 2023 and 2024 sequences shared the K276E/K276R substitutions, while the 2024 sequence uniquely exhibited the L86X substitution (Supplementary Figure 4C). In the neuraminidase (NA) gene, the 2023 sequence displayed the D346G substitution, whereas the 2024 sequence showed additional substitutions, including M51I, I469T, R400K, S44X, and R150H (Figure 4D).

## Discussion

In this study we have investigated the influenza strains circulating in the Western Province of Sri Lanka from 2023 to mid-2024, providing detailed analysis of the circulating clades of Influenza A. The frequency of both influenza A and influenza B was predominantly seen in children <10 years of age while SARS-CoV-2 infection was seen in adults >60 years of age. Many studies have shown that individuals at extremes of age, including children, have shown to be vulnerable to be hospitalized due to influenza [14]. However, in our cohort SARS-CoV-2 accounted for most infections in those >60 years of age (22.7%) compared to 9.1% of infections due to influenza A. Sri Lanka did not receive any COVID-19 vaccines as booster since 2022 [15] and therefore, elderly individuals and those with comorbidities are at increased risk of hospitalization due to COVID-19, possibly due to waning of immunity. In our cohort, 4 individuals had co-infection with influenza A and B, while one patient had co-infection with influenza and SARS-CoV-2. Co-infections with influenza A and B have been previously reported [16–18], and have shown to associate with a worse disease outcome [17]. We also reported one patient with co-infection with influenza and SAR-CoV-2, which has also previously been reported [19]. Due to the limited sample size, we could not determine if co-infections were associated with worse disease outcomes.

Seasonal influenza outbreaks usually coinciding with the monsoon season in tropical and subtropical regions [7]. As reported in the Global Influenza Surveillance and Response System, of the WHO, a similar pattern is observed in the Western Province, Sri Lanka, where there are two influenza A seasons, which are from November to January and again from April to June [20]. During early 2023, the predominant influenza A subtype was H1N1, which was replaced by H3N2 as the predominant subtype by June 2023. In 2024, again H1N1 became the predominant subtype. These changes are consistent with the changes in the influenza A subtypes in India and Nepal, but different to the changes in subtypes seen in Bangladesh, Thailand and Bhutan [20].

In our study, all H1N1 sequences from 2023 and 2024 were classified within the 6B.1A.5a.2a clade. Our sequences, characterized by substitutions I418V and v47i, were placed within the C.1 subclade and its associated subclusters [21]. Similar H1N1 strains dominated in Southeast Asia, the Middle East, Africa, Central America, and parts of Europe [21]. The 5a.2a.1 clade, which has become more prevalent in the United States, Caribbean, Japan, and several European countries, marked by mutations like P137S and K142R, has significantly diverged from the 5a.2a clade in 2023 [21]. This antigenic drift resulted in reduced effectiveness of the 5a.2a-based vaccine, represented by the A/Sydney/5/2021 strain, leading the WHO to update the vaccine to target the 5a.2a.1 clade for the 2024 season, now represented by A/Wisconsin/67/2022 and A/Victoria/4897/2022 [13]. Our influenza A H1N1 strains in 2024 had the additional mutations I96T, T120A, A139D, G339X, K156X, and T278S. Although the positions and the function of these genes which carried these mutations are known, the significance of these mutations in relation to vaccine efficacy or virulence of the virus is not known. Therefore, it would be important to continue surveillance to understand if the influenza vaccine containing the strains of 5a.2a.1 provides protection against both 5a.2a and 5a.2a.1 viruses, currently circulating in Sri Lanka.

Our phylogenetic analysis of the A(H3N2) HA gene sequences revealed the circulation of the 3C.2a1b.2a.2a.1b clade in 2023 and the 3C.2a1b.2a.2a.3a.1 clade in 2024. In 2023, the 3C.2a1b.2a.2 subclade, characterized by mutations such as I140K and K276R, was the most prevalence strain globally [20]. However, by 2024, the 3C.2a1b.2a.3 subclade, particularly the 2a.3a.1 lineage, emerged as the dominant strain [20, 21]. Our 2024 sequences aligned with the .2a.3a.1 clade (clade J), marked by mutations such as K276E and V223I [21]. These changes led to significant antigenic drift, reducing the efficacy of the A/Darwin/9/2021-based vaccine, which was updated for 2024 including the A/Thailand/8/2022 and A/Massachusetts/18/ strains[21]. Our 2024 strains have additional mutations such as N122D and K276E. Although the effect of these mutations on vaccine efficacy is not clear, it would be important to continue surveillance to detect further emerging influenza strains.

In summary, in this study we have characterized the influenza A strains that circulated in Sri Lanka over a period of 18 months. We found that all H1N1 sequences from 2023 and 2024 were classified within the clade 6B.1A.5a.2a clade, while the H3N2 sequences in 2023 were assigned to clade 3C.2a1b.2a.2a.1b and the 2024 strains to clade 3C.2a1b.2a.2a.3a.1. As the Sri Lankan strains also had certain mutations of unknown significance, it would be important to continue detailed surveillance of the influenza strains in Sri Lanka to choose the most suitable vaccines for the population and the timing of vaccine administration.

## Data Availability

All data produced in the present work are contained in the manuscript.

## Acknowledgments

We are grateful to the NIH, USA (grant number 5U01AI151788-02) and the UK Medical Research Council for funding, also thankful to Asia Pathogen Genomic Initiative for providing Influenza sequencing primers.

## Notes

### Competing Interest Statement

The authors have declared no competing interest.

