## Supplementary figures for "Genomic Surveillance and Evolutionary Dynamics of Influenza A Virus in Sri Lanka"

| H1N1 HA gene | H1N1 NA gene | H3N2 HA | H3N2 NA |
| --- | --- | --- | --- |
| EPI_ISL_19288002 | EPI_ISL_19288002 | EPI_ISL_19287905 | EPI_ISL_19287905 |
| EPI_ISL_19288000 | EPI_ISL_19290528 | EPI_ISL_19287998 | EPI_ISL_18667508 |
| EPI_ISL_19290528 | EPI_ISL_19288000 | EPI_ISL_18667554 | EPI_ISL_19287998 |
| EPI_ISL_19288001 | EPI_ISL_19288001 |  | EPI_ISL_18667554 |
| EPI_ISL_18651856 | EPI_ISL_18651856 |  |  |
| EPI_ISL_18667474 | EPI_ISL_18667506 |  |  |
| EPI_ISL_18667506 | EPI_ISL_18667474 |  |  |
| EPI_ISL_18667468 | EPI_ISL_18667468 |  |  |
| EPI_ISL_18667405 | EPI_ISL_18667404 |  |  |
| EPI_ISL_19290526 | EPI_ISL_18667469 |  |  |
| EPI_ISL_18667557 | EPI_ISL_18667405 |  |  |
| EPI_ISL_19290527 | EPI_ISL_19290526 |  |  |
| EPI_ISL_18667558 | EPI_ISL_18667556 |  |  |
| EPI_ISL_19290525 | EPI_ISL_19290527 |  |  |
|  | EPI_ISL_18667557 |  |  |
|  | EPI_ISL_18667558 |  |  |
|  | EPI_ISL_19290525 |  |  |

**Supplementary data**

Supplementary table 1: Accession numbers of HA and NA sequences of H1N1 and H3N2 included in the analysis


Supplementary table 2: Positions of the mutations and gene functions in 2024 H1N1 sequences from Sri Lanka

| Mutation | Impact | Protein structure |
| --- | --- | --- |
| I96T | Viral oligomerization interfaces, binding small ligand(s), a T-cell epitope presented by MHC molecules, antibody recognition sites | Helix |
| T120A | Binding small ligand(s), viral oligomerization interfaces, antibody recognition sites | Beta strand |
| A139D | Antigenic drift / escape mutant, viral oligomerization interfaces, binding small ligand(s), antibody recognition sites | Adjacent to turn |
| G339X | Viral oligomerization interfaces, binding small ligand(s), antibody recognition sites | NA |
| K156X | Antibody recognition sites, binding small ligand(s), viral oligomerization interfaces | Prior to beta strand, inbetween two beta strands |
| T278S | Binding small ligand(s), viral oligomerization interfaces | Inbetween two beta strands |

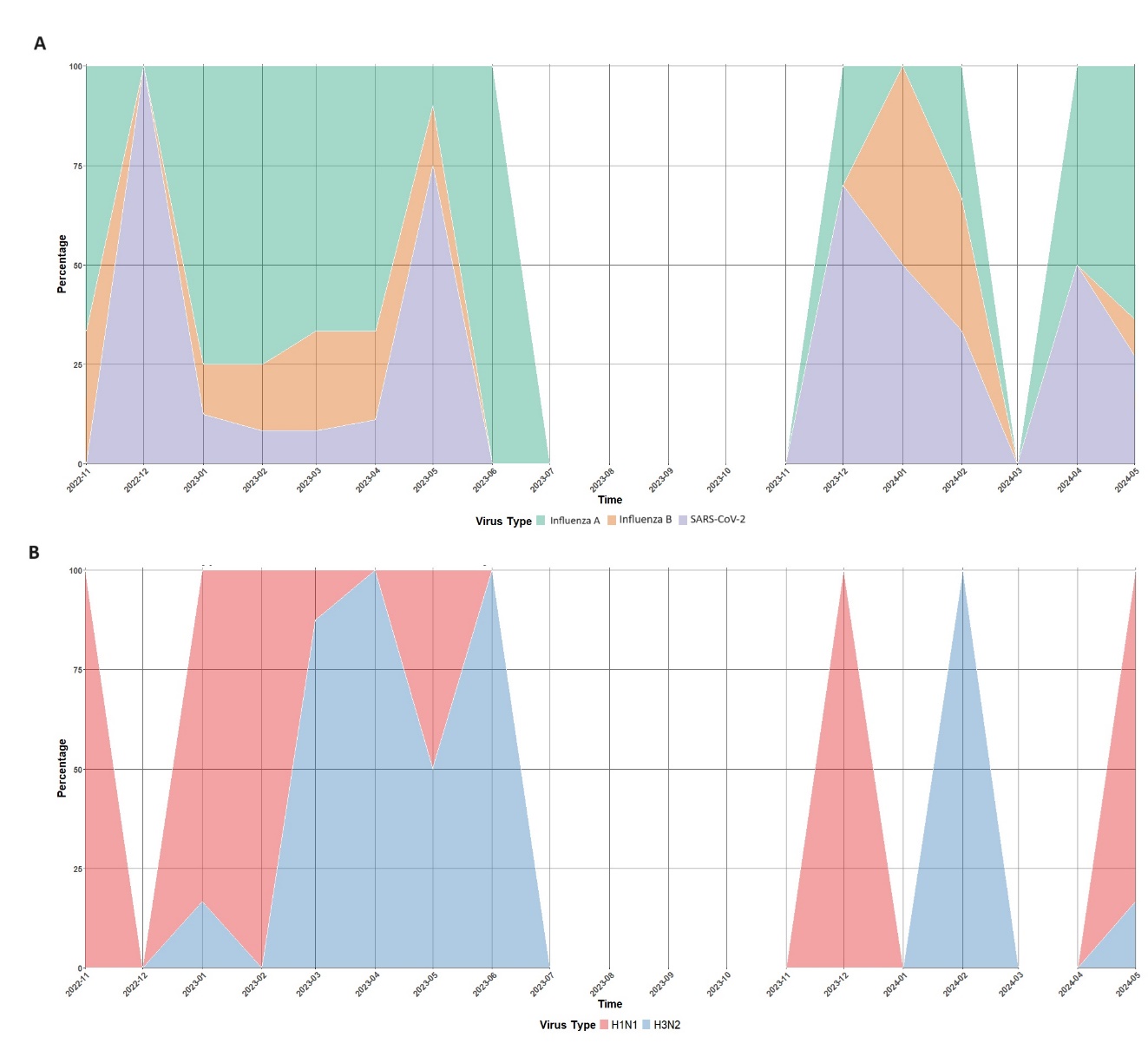

Supplementary figure 1: (A) Monthly trend of Influenza A, Influenza B, and SARS-CoV-2 from November 2022 to May 2024. (B) Influenza Subtype distribution from November 2022 to May 2024

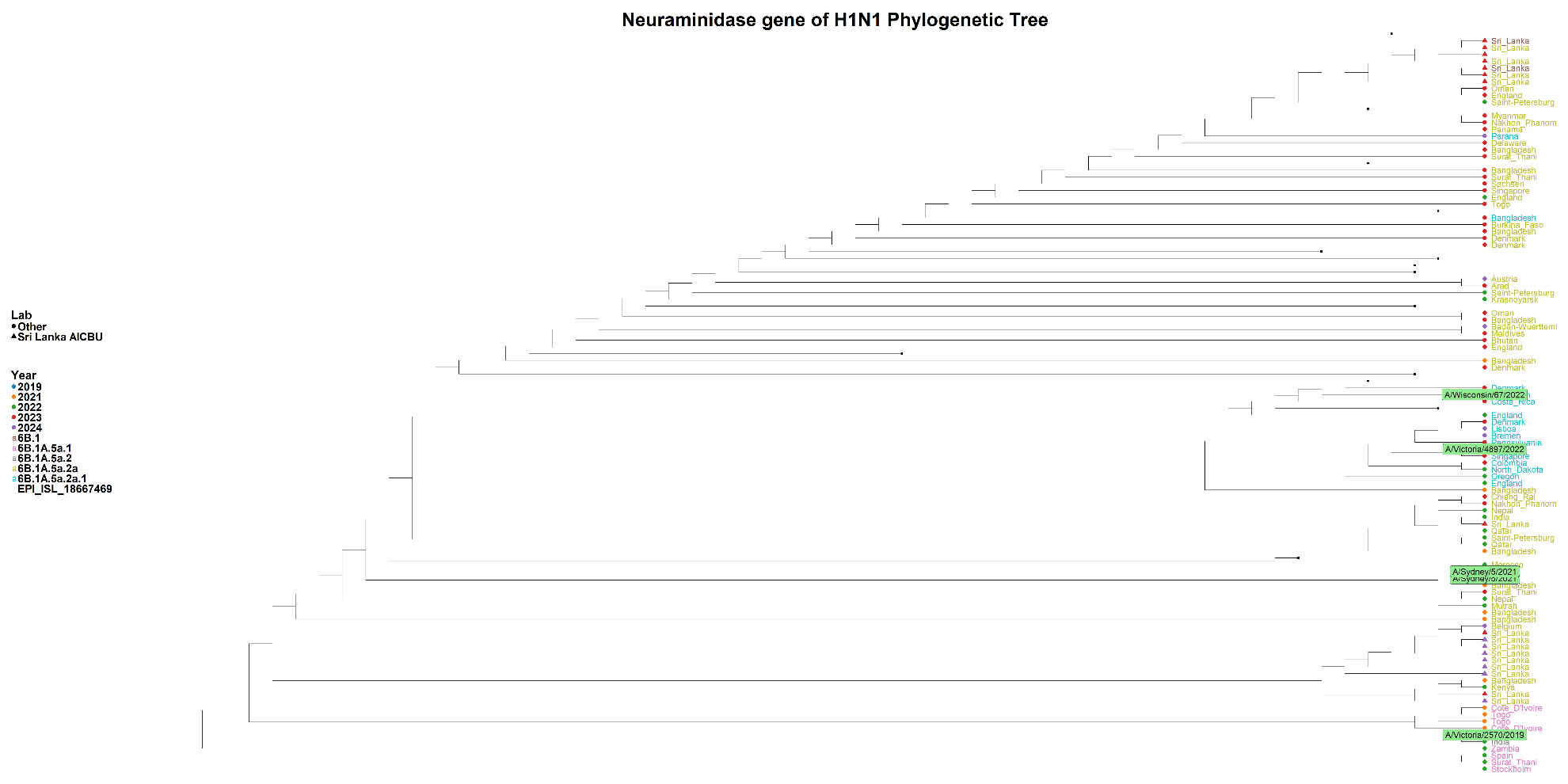

Supplementary figure 2: Phylogenetic tree of the H1N1 NA gene (Sequences generated in the AICBU lab are represented by triangles, while other sequences are depicted as circles)

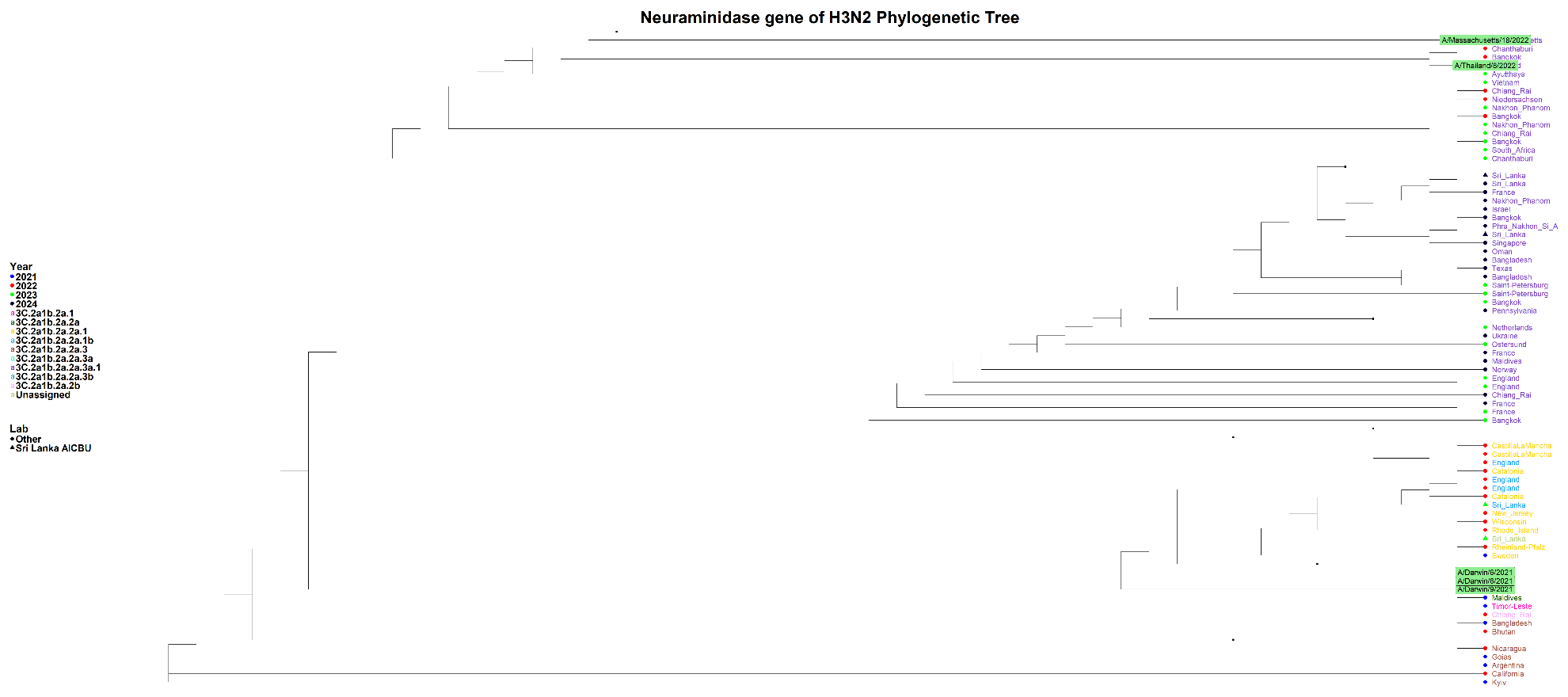

Supplementary figure 3: Phylogenetic tree of the H3N2 NA gene (Sequences generated in the AICBU lab are represented by triangles, while other sequences are depicted as circles)

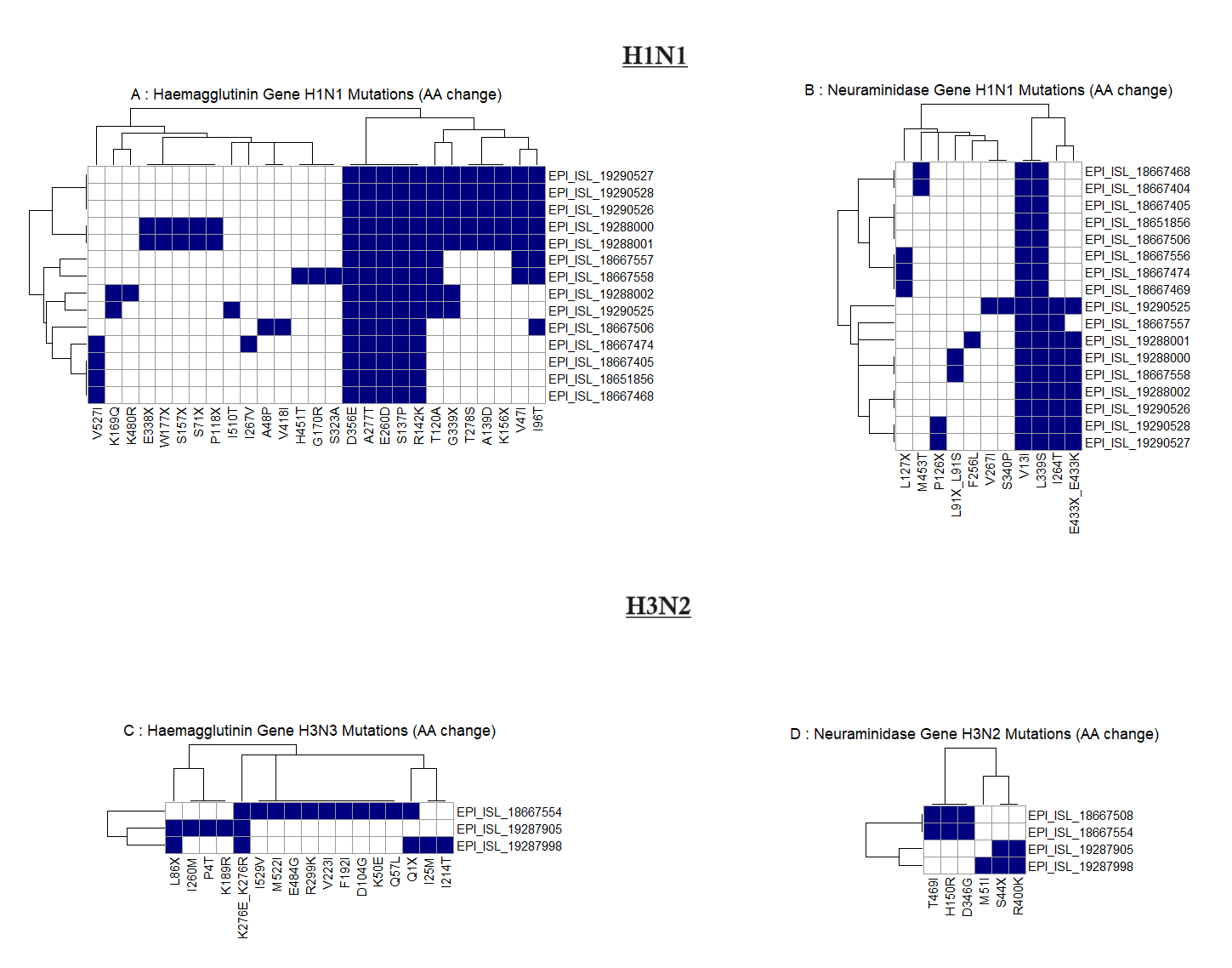

Supplementary figure 4: Heatmap of amino acid substitutions in the Hemagglutinin (HA) and Neuraminidase (NA) genes of Influenza A H1N1 and H3N2 viruses. The reference strain A/Wisconsin/67/2022 was used for H1N1, and A/Massachusetts/18/2022 was used for H3N2. (Panels A and B illustrate mutations in the HA and NA genes of H1N1, respectively, while Panels C and D depict mutations in the HA and NA genes of H3N2. Each row corresponds to an individual virus sequence, identified by its GISAID EPI_ISL accession number, and each column represents specific amino acid positions where mutations have been identified. Blue squares indicate the presence of a mutation, while white squares indicate its absence.)
